# Clinical outcome evaluations and CBT response prediction in Myotonic Dystrophy

**DOI:** 10.1101/2021.02.25.21252140

**Authors:** Daniël van As, Kees Okkersen, Guillaume Bassez, Benedikt Schoser, Hanns Lochmüller, Jeffrey C. Glennon, Hans Knoop, Baziel G.M. van Engelen, Peter A.C. ’t Hoen, on behalf of the OPTIMISTIC consortium

## Abstract

**Background:** The European OPTIMISTIC clinical trial has demonstrated a significant, yet heterogenous effect of Cognitive Behavioural Therapy (CBT) for Myotonic Dystrophy type 1 (DM1) patients. One of its remaining aims was the assessment of efficacy and adequacy of clinical outcome measures, including the relatively novel primary trial outcome, the DM1-Activ-c questionnaire.

**Objectives:** Assessment of the relationship between the Rasch-built DM1-Activ-c questionnaire and 26 commonly used clinical outcome measurements. Identification of variables associated with CBT response in DM1 patients.

**Methods:** Retrospective analysis of the to date largest clinical trial in DM1 (OPTIMISTIC), comprising of 255 genetically confirmed DM1 patients randomized to either standard care or CBT with optionally graded exercise therapy. Correlations of 27 different outcome measures were calculated at baseline (cross-sectional) and of their respective intervention induced changes (longitudinal). Bootstrap enhanced Elastic-Net (BeEN) regression was validated and implemented to select variables associated with CBT response.

**Results:** In cross-sectional data, DM1-Activ-c correlated significantly with the majority of other outcome measures, including Six Minute Walk Test and Myotonic Dystrophy Health Index. Fewer and weaker significant longitudinal correlations were observed. Nine variables potentially associated with CBT response were identified, including measures of disease severity, executive cognitive functioning and perceived social support.

**Conclusions:** The DM1-Activ-c questionnaire appears to be a well suited cross-sectional instrument to assess a variety of clinically relevant dimensions in DM1. Yet, apathy and experienced social support measures were less well captured. CBT response was heterogenous, requiring careful selection of outcome measures for different disease aspects.

## Introduction

Myotonic dystrophy type 1 (DM1) is a genetic, complex multisystem progressive disorder caused by a trinucleotide repeat expansion in the dystrophia myotonica protein kinase (*DMPK*) gene(1,2). DM1 is a disorder associated with a high emotional, social and economic burden and loss of quality of life(3–6). The unmet clinical need in DM1 is high, as there is no cure and options for symptomatic treatment are limited. As for any rare disease, the design of properly powered clinical trials in DM1 is difficult and calls for sensitive and robust outcome measures(7,8).

The European OPTIMISTIC project comprised of a 4-country prospective clinical trial that included 255 genetically proven, adult DM1 patients(9,10). They were randomized 1:1 to standard care versus standard care with a behavioural intervention that comprised of cognitive behavioural therapy (CBT), optionally combined with graded exercise. Besides assessment of efficacy of the behavioural intervention, the results of which have been published elsewhere(10), one of the aims of OPTIMISTIC was the identification of outcome measures for assessment of efficacy and adequacy of clinical response. Identification of appropriate outcome measures may aid in the design and conduction of future (drug) trials. Therefore, many patient-reported outcome measures (PROMs), clinical tests and objective assessments were collected during the trial. The primary outcome was the relatively new DM1-Activ-c scale, a DM1-specific, 25-item Rasch-built patient reported outcome measure for (capacity for) activity and social participation that yields a 0-100 interval score(11,12).

As the OPTIMISTIC study was the first occasion on which the DM1-activ-c was used in a prospective clinical trial, we were interested in how it relates to other outcome measures that have been used in previous DM1 trials or clinical trials in other neuromuscular diseases. Additionally, we were interested in the individual changes of DM1-Activ-c in the intervention group, as significant heterogeneity in the response to CBT was observed(10). In future studies and in clinical practice, identification of those individuals likely to benefit from the intervention (i.e. prediction), would be highly valuable. We therefore studied whether we could identify baseline variables associated with a beneficial treatment response, as measured by means of DM1-Activ-c changes over the course of 10 months. To this end, we validated and implemented a machine learning based regression solution (Bootstrap enhanced Elastic-Net)(13–15).

In summary, our objectives were to evaluate both cross-sectional (1) and longitudinal, intervention driven (2), correlations among patient reported outcome measures, clinical tests and accelerometery data used in the OPTIMISTIC clinical trial. Furthermore, Bootstrap enhanced Elastic-Net has been validated (3) and used to identify predictors of a positive cognitive behavioural therapy response for DM1 patients (4).

## Methods

### Dataset

All data used to perform this study were collected during the OPTIMISTIC trial. Trial specific information, including ethical committee approval and methodological details have been published elsewhere(10). In short, 255 genetically proven, adult, ambulant, DM1 patients that were severely fatigued (CIS-fatigue score ≥ 35) from four clinical sites [Paris, France (n=71, 28%); Munich, Germany (n=66, 26%); Newcastle, UK (n=52, 20%) and Nijmegen, Netherlands (n=66, 26%)], were included after providing written informed consent. Participants in the OPTIMISTIC study were on average 45.6 years old, with an estimated age at onset of 25.4 years; 117 participants (46%) were female. Participants were randomized 1:1 to either standard care (n=127) or standard care with a behavioural intervention (n=128). The latter comprised of cognitive behavioural therapy (CBT), optionally combined with graded exercise. CBT was delivered by trained psychologists or psychotherapists and based on a specifically developed treatment manual; it was tailored to the individual patient on the basis of screening questionnaires and an intake session with the therapists. CBT was given over a 10-month period and was front-loaded with the majority of sessions given within the first five months. The average number of sessions was 10, with each session scheduled for a duration of 40-60 minutes. Clinical assessments were obtained at the start of the trial (referred to as baseline measurements) and after 5, 10 and 16 months. Summaries of the variables and clinical assessments that were obtained during the OPTIMISTIC trial are given in table 1 and 2. Our analyses focus on the difference after 10 months, as it was the primary clinical trial endpoint where the largest effect of the CBT intervention has been observed. The trial was powered for both DM1-Activ-c, the primary outcome measure, and for the six-minute walk test (6MWT), a measure of exercise capacity. Researches whishing to access the OPTIMISTIC trial data should contact BGMvE (10).

**Table 1:** Overview of OPTIMISTIC measurements(10)

|  | Measurement | Score Range | Direction* |
| --- | --- | --- | --- |
| **Primary outcome** |  |  |  |
| DM1-Activ-c(12) | Capacity of Activity and Social Participation | 0 – 100 | H |
| **Secondary outcomes** |  |  |  |
| Six-minute walk test (6MWT)(29) | Exercise capacity | 0 – ∞ | H |
| BORG Scale (PreBORG)(30) | Perceived exertion | 0 – 10 | L |
| Myotonic Dystrophy Health Index (MDHI)(31) | Disease impact | 0 – 100 | L |
| Fatigue and Daytime Sleepiness Scale (FDSS)(32) | Experienced fatigue and sleepiness | 0 – 100 | L |
| Checklist Individual Strength – subscale fatigue (CISFatigue)(33) | Experienced fatigue | 8 – 56 | L |
| Accelerometry – Euclidian Norm Minus One (ENMO); divided in least, mean and maximum 5 hours of activity (resp. L5ENMO, MeanENMO, M5ENMO)(34) | Activity | 0 – ∞ | H |
| Individualized Neuromuscular Quality of Life Questionnaire – domain quality of life (INQoL)(35) | Quality of life / health status | 0 – 100% | L |
| Beck Depression Inventory – fast screen (BDIFS)(36) | Depression | 0 – 21 | L |
| Apathy Evaluation Scale – clinical version (AESc)(37) | Apathy | 18 – 72 | L |
| Stroop colour-word interference score (Stroop)(38)** | Executive cognitive functioning | 0 – ∞ | H |
| **Exploratory outcome measures** |  |  |  |
| Muscle Impairment Rating Scale (MIRS)(39) | Progression in muscular impairment | 1 – 5 | L |
| Trail Making Test (TMT)***(40) | Executive cognitive functioning | 0 – ∞ | L |
| McGILL Pain Questionnaire(41)<br>– short version (McGILLPain) | Severity of experienced pain during last 7 days | 0 – 100 | L |
| Adult Social Behavioral Questionnaire (ASBQ)(42) | Social behaviour | 44 – 132 | L |
| Social Support(43)<br>– Discrepancies (SSLD) | Combination score of perceived lack or surplus of social support | 34 – 102 | L |
| Social Support(43)<br>– Negative Interactions (SSLN) | Negative interactions | 7 – 28 | L |
| Social Support(43)<br>– Interactions (SSLI) | Experienced social support | 34 – 136 | H |
| Jacobsen Fatigue Catastrophizing Scale (JFCS)(44) | Attitudes and perception of fatigue | 10 – 50 | L |
| Illness Cognition Questionnaire(45)<br>– subscale acceptance (ICQ) | Degree of coping/accepting disease | 6 – 24 | H |
| Illness Management Questionnaire (IMQ)(46) | Dealing with chronic disease | 9 – 54 | L |
| Self-Efficacy Scale 28 (SES28)(47) | Attitude towards fatigue | 7 – 28 | H |
| Caregiver Strain Index (CSI)(48) | Strain on caregiver | 0 – 13 | L |
| Apathy evaluation scale(49)<br>– informant version (AESI) | Apathy | 18 – 72 | L |
| Checklist Individual Strength(33)<br>– subscale activity (CISActivity) | Reduction of activities | 3 – 21 | L |
| Sickness Impact Profile (SIPScore)(50)<br>– subscale Sleep & Rest | Quality of sleep and rest at baseline | 0 – 499 | L |

\* H: Higher scores beneficial; L: Lower scores beneficial
\*\* Calculated by dividing the accuracy-speed trade-off of Stroop III by the accuracy-speed trade-off of Stroop II
\*\*\* Calculated by dividing TMT-B completion time by TMT-A completion time

**Table 2:**
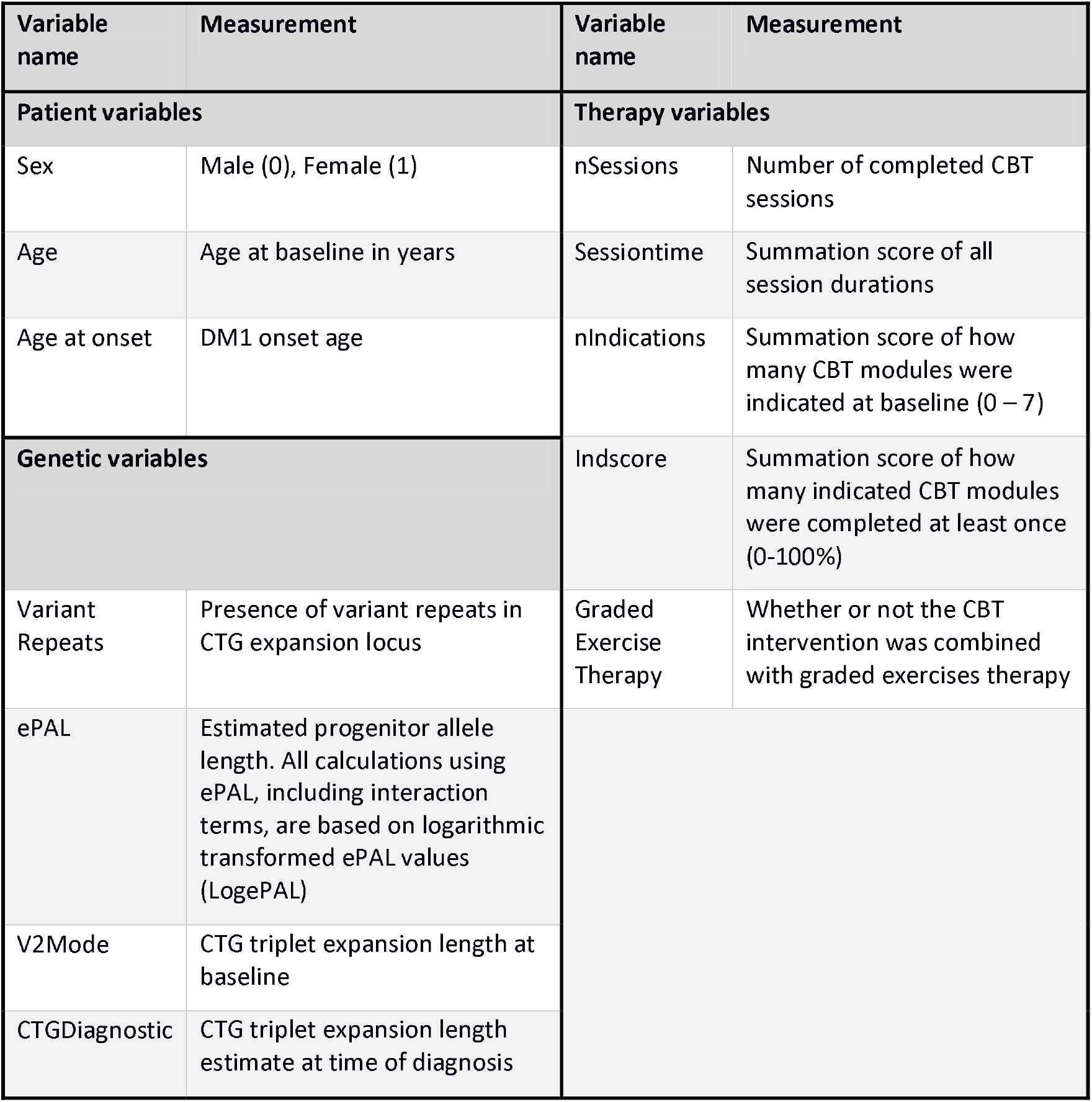
Patient specific variables.

## Statistical analyses

### Correlation analysis

We performed two correlation analyses: (1) correlations between baseline variables as collected in OPTIMISTIC (hereafter: cross-sectional correlations) and (2) correlations between the absolute, intervention driven changes in variables from t=0 to t=10 months (hereafter: longitudinal correlations). We calculated cross-sectional correlations based on the entire study cohort (n=255) and calculated the longitudinal correlations based on the intervention group of the study (n=128). All 27 variables used for these analyses are listed in table 1.

Analysis of the variable distributions using the Shapiro-Wilk’s test for normality revealed that the majority of the variables deviated significantly from the normal distribution. Screening for outliers was based on the Interquartile Range Rule (Q1 – 2*IQR and Q3 + 2*IQR as respective lower and upper boundaries), with flagged outliers being individually confirmed using distribution plots. At baseline 6 outliers were removed, and for the delta-values 18 outliers were removed.

Pairwise correlations were calculated based on the non-parametric Spearman’s rank methodology using the function ‘corr.test’ of the R package ‘psych’ (v 1.8.12)(16). Using this function, statistical significance was calculated by a formula based conversion of the individual correlation scores to a t-statistic. Adjustment for multiple testing was done by setting the false discovery rate (FDR) to p = 0.05 using the Benjamini-Hochberg procedure. Statistically significant results were visualized using the R package ‘corrplot’ (v 0.84), with nominal significant results under and adjusted significant results above the diagonal(17).

### Regression analyses

The second part of our study aimed at the identification of baseline variables associated with CBT intervention effect, defined as 10-month change in DM1-Activ-c scores. Statistical modelling was challenged by both the high number of baseline predictors in relation to the patient cohort size (multidimensionality) as well as the significant correlations among the predictors (multicollinearity). These challenges can result in unstable regression coefficients with high standard errors when using regular linear regression solutions(14,18). Additionally, a clinically useful prediction system is limited to a subset of important predictors, making a variable selection approach necessary. The machine learning based Bootstrap enhanced Elastic-Net (BeEN) regression approach is a powerful variable selection tool that has been shown to overcome the limitations of high dimensionality and multicollinearity(13–15). Using this approach, predictors are selected independently for each bootstrap distribution based on Elastic-Net regression. Subsequently, variable inclusion probabilities (VIP) are calculated based on the percentage of bootstraps in which the predictors were selected(14,15). VIP’s ranging between 50-70% yield sensitive, yet adequately conservative predictor selection results(14,15,18).

### Regression framework validation

To validate Bootstrap enhanced Elastic-Net (BeEN) regression, it was first used to reproduce the published regression model of baseline DM1-Activ-c scores. Cumming et al. used backwards stepwise regression (BSR) on the OPTIMISTIC data to predict baseline DM1-Activ-c scores(19):

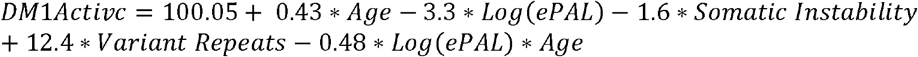

We generated an exact reproduction of this model using the stepAIC function of the R package ‘MASS’ (v 7.3.51.5) on 246 patients of the OPTIMISTIC datasets(20). Next, we implemented BeEN regression using the R packages ‘glmnet’ (v 2.0.16) and ‘boot’ (v 1.3.20) by using the cv.glmnet function with alpha=0.5 and 5 fold cross validation on 5000 different bootstrap distributions(21,22). Per predictor, its variable inclusion probability (VIP) was calculated based on the percentage of its non-zero coefficients among the 5000 bootstrap distributions. We selected VIPs greater or equal to 60% and subsequently used them to fit a linear regression model on the same dataset. If an interaction term has been selected, the respective main effects were selected as well. As a measure of linear regression model performance, we report the adjusted R-squared (a.Rsq) as well as the Root Mean Squared Error (RMSE) for both models.

To study the external validity and the impact of overfitting, we split the dataset in training (n=184, 75% cases) and testing (n=62, 25% cases) subsets. We used the training set to both selected variables using either BSR or BeEN regression and subsequently build ordinary least squares models. We calculated model specific out-of-sample (OS) statistical performance measures (a.Rsq and RMSE) based on the prediction results of the testing set. To account for randomness based on the training / testing splits this was repeated 10 times. Through selection of identical cohort splits for both regression approaches, we assured comparability. We report mean VIPs and statistical performance measures for both approaches, in addition to box-blots of sampling dependent statistical performance distributions.

As our proposed main analysis was restricted to the intervention group, it was also of interest to study the impact of reducing the patient cohort by roughly 50% on variable inclusion frequencies as well as on statistical performance measures of both models. Therefore, the aforementioned analyses were repeated for the intervention group (n=126, 10 subsets distinct subsets with each n=94/32).

### Variable selection for delta-DM1-Activ-c prediction

CBT treatment response was defined as changes in the primary OPTIMISTIC research outcome DM1-Activ-c between 10 months and baseline, as expressed in delta values (dDM1-Activ-c). All variables listed in table 1 and 2 were considered as independent variables, resulting in an initial set of 40 distinct potential predictors. This analysis was limited to the patients of the intervention cohort (n=128). Data from 22 patients were excluded from the analysis for the following reasons: missing essential treatment related information (n=7), missing DM1-Activ-c scores at baseline or at t=10 months (n=11), maximum baseline DM1-Activ-c scores (n=4). Patients with maximum DM1-Activ-c baseline values were removed as those cases cannot be used to identify variables associated with positive treatment response. We identified and removed 2 outliers by combining the IQR rule (Q1 – 2*IQR and Q3 + 2*IQR as respective lower and upper boundaries) with visual inspection.

Baseline variables with more than 25% missing values among all cases were excluded (CSI, AESI, CTGDiagnostic). We imputed three missing values regarding CBT therapy session time of two patients by averaging the session times of their respective other sessions. The remaining missing values were imputed using the ‘Multiple Imputation by Chained Equations’ (MICE) algorithm (v. 3.4.0)(23). A total of 10 different imputed datasets were generated, each using 50 iterations for predictive mean matching. The impact of imputation was mainly studies by its influence on variable inclusion probability (VIPs).

Each imputed dataset was used to fit Elastic-Net regression on 5000 bootstrap distributions using the R functions ‘boot’ and ‘cv.glmnet’ with alpha=0.5 and 5 fold cross validation(21,22). Subsequently, we calculated VIPs for each variable based on the percentage of their non-zero coefficients among the 5000 bootstrap distributions. Imputation independent predictors were obtained by calculating mean VIP’s among all imputed datasets. Predictors with a mean VIP exceeding 60% were selected and used to fit a linear model on 80 unimputed intervention cases without missing relevant values. After removal of one outlier (n=79), all assumptions of linear regression as assessed by the R package ‘gvlma’ (v 1.0.0.3) were met(24). Unstandardized regression coefficients as well as their error terms are reported. In order to be able to distinguish variables that mostly predict therapy response versus natural history, we implemented the same regression framework in the control group, excluding treatment related variables.

All statistical analyses were done using the programming language R (version 3.5.1). All R scripts used to generate the results are available on <u>GitHub</u>.

## Results

The clinical trial OPTIMISTIC investigated the value of cognitive behavioural therapy (CBT) with optional graded exercise therapy for patients with genetically confirmed DM1. The primary research outcome was measured with the Rasch-built DM1-Activ-c questionnaire. In addition to the primary outcome measure, 12 secondary outcome measures were used, as well as 15 exploratory outcome measures. A comprehensive overview of all outcome measures, clinical tests and accelerometery measurements is summarized in Table 1. Patient specific variables such as patient characteristics, genetic variables and CBT related information are summarized Table 2.

### Correlation analyses

The first objective of this study was the identification of interrelations between the different outcome measures used in the OPTIMISTIC trial, as significant interrelations may reveal redundancy. This could potentially limit the number of outcome measures used in future trials, thereby decreasing patient burden while also improving patient adherence and study efficiency. Interrelations between outcome measures were studied by calculating pairwise correlations at baseline (cross-sectional correlations), as well as their changes after 10 months in response to CBT (longitudinal changes).

### Cross-sectional correlations

The DM1-Activ-c scores correlated significantly with 17 other variables (BH adjusted p-value (p-adj) < 0.05; Fig 1A). DM1-Activ-c scores demonstrated the strongest correlations with 6MWT (rho = 0.66, p-adj < 0.001) and the Myotonic Dystrophy Health Index (MDHI, rho = -0.63, p-adj < 0.001). In general, many of the variables were correlated, with the strongest correlations between the designated primary and secondary outcome measures in the OPTIMISTIC trial (Fig 1 and Table 1). Notable exceptions were scores for apathy, experienced social support and strain on caregivers.

**Figure 1:**
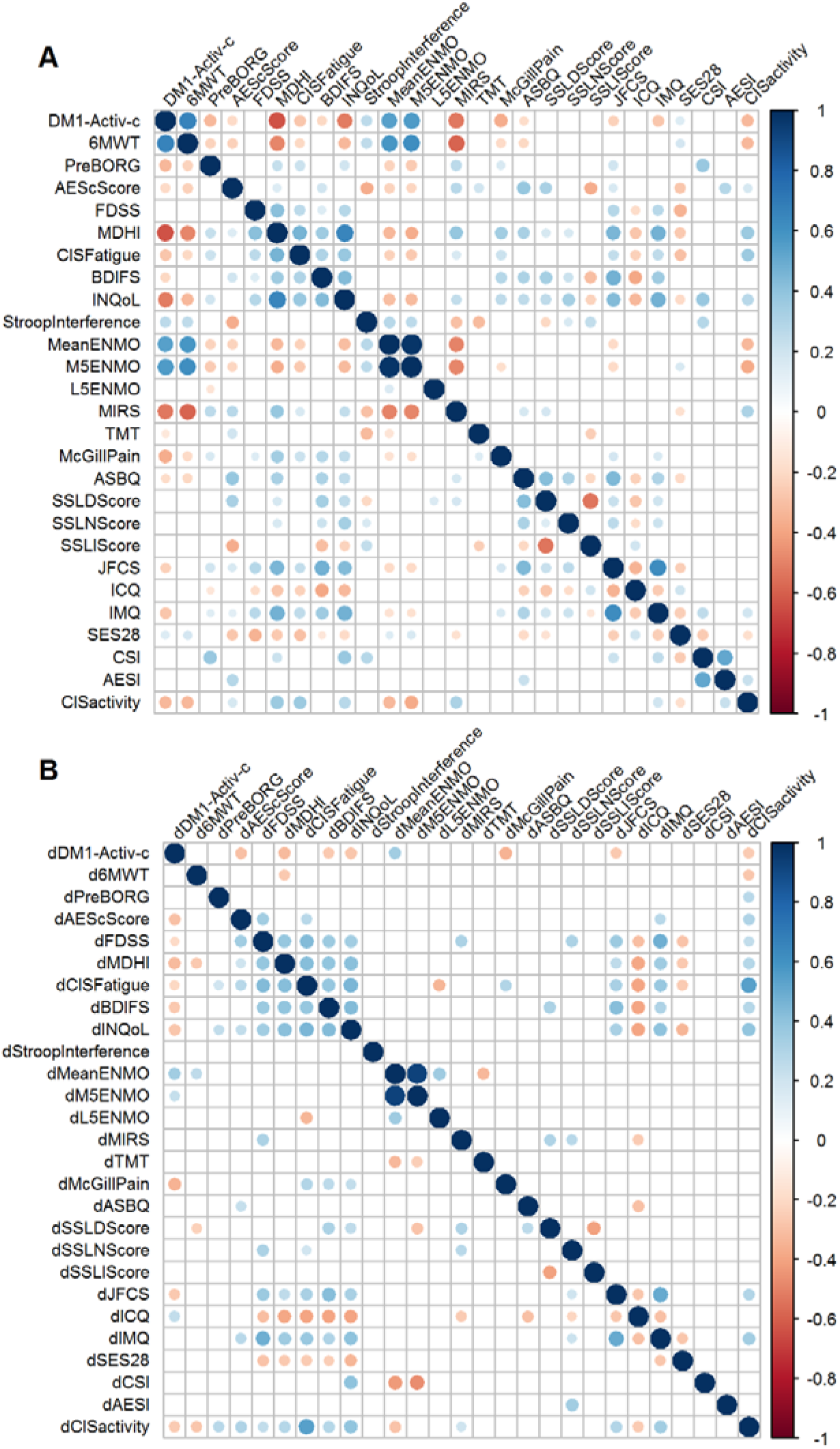
Correlograms of OPTIMISTIC baseline and delta-value measurements. Spearman rank correlations of OPTIMISTIC measurements at baseline (N=255; panel **A**) and their delta-values (difference in intervention group between the 10 months and baseline evaluation; N=128; panel **B**). Significance was calculated using t-tests in a pairwise manner. Only nominally significant (p < 0.05, below the diagonal) pairwise correlations and pairwise correlations that are significant after multiple testing correction (Benjamini-Hochberg adjusted p<0.05, above the diagonal) are shown as circles. The size of the circle represents the magnitude of the Spearman rho correlation, the color represents the direction.

### Longitudinal correlations

Overall, we observed fewer and weaker correlations in the longitudinal correlation analysis in comparison to the cross-sectional analysis (Fig 1B). Yet, the majority of the BH-adjusted significant baseline correlations were also found for the delta-value correlations, defined as the score at t = 10 months minus t = 0 months. The lower number of subjects (only 128 subjects in the intervention group) was not the only reason for the lower significance; the correlations were also weaker (absolute rho) in comparison to the cross-sectional correlations. Delta-DM1-Activ-c (dDM1-Activ-c) correlated significant with 8 other delta scores (BH adjusted p-value (p-adj) < 0.05). The strongest correlations of dDM1-Activ-c we observed were with dMDHI (rho = -0.32, p-adj = 0.01) and the accelerometer measure dMeanENMO (rho = 0.34, p-adj = 0.03). Notably, we did not observe a longitudinal correlation between DM1-Activ-c and 6MWT, despite their moderately-strong cross-sectional correlation (rho = 0.14, p-adj = 0.38). Nevertheless, all significant cross-sectional and longitudinal correlations were in the expected direction: DM1-Activ-c correlated positive with measurements in which patients benefit from a higher score and negatively with measurements in which negative scores are beneficial.

### Regression analyses

Although the OPTIMISTIC clinical trial demonstrated positive effects of CBT on activity, participation and exercise capacity of DM1 patients, substantial heterogeneity in clinical responses have been observed. One important objective of this study was therefore the identification of baseline measurements and patient characteristics that are associated with CBT treatment response. Treatment response was defined as the 10 month changes of DM1-Activ-c scores in the intervention group, expressed in delta-values. All variables listed in Table 1 and 2 were considered as potential predictors. Given the large number of significantly correlating predictors, conventional regression solutions could not be used as they are likely to yield unstable results(14,18). We therefore proposed the use of the machine learning based Bootstrap enhanced Elastic-Net (BeEN) regression. This algorithm has been shown to overcome these limitations and is furthermore able to select predictors(13). Among a large number of bootstrap distributions, predictors are chosen based on how frequently they have been independently selected(14,15). This frequency is called variable inclusion probability (VIP) and predictors with a high VIP are considered to be important predictors. To validate this methodology, we first compared it to the more commonly used Backwards Stepwise Regression (BSR) approach that has been used to predict baseline DM1-Activ-c scores(19).

### Regression framework validation

Both, BeEN regression and its comparator BSR, selected the variables ePAL, Variant Repeats, Age and the interaction term Age*ePAL as predictors for baseline DM1-Activ-c (Table 3). The BSR approach additionally identified somatic instability (SI) as a meaningful predictor. The resulting regression models show similar, yet low performance measures for both a-Rsq (0.23, 0.22) and RMSE (15.06, 15.15) for respectively the BSR and BeEN regression models (Table 4).

**Table 3:** Comparison of predictor inclusion frequencies (%) between BSR and BeEN based regression.

| Cohort* | Cohort sizes** | Method*** | ePAL | VR | Age | SI | Sex | Age*ePAL | ePAL*SI | Age*SI | Age*ePAL*SI |
| --- | --- | --- | --- | --- | --- | --- | --- | --- | --- | --- | --- |
| FC | 246 | BSR | 100 | 100 | 100 | 100 | 0 | 100 | 0 | 0 | 0 |
| FC | 246 | BeEN | 100 | 100 | 100 | 0 | 0 | 100 | 0 | 0 | 0 |
| FC | 184 / 62 | BSR | 100 | 100 | 100 | 70 | 0 | 80 | 50 | 50 | 50 |
| FC | 184 / 62 | BeEN | 100 | 70 | 100 | 0 | 0 | 100 | 0 | 0 | 0 |
| IC | 126 | BSR | 100 | 0 | 100 | 100 | 0 | 0 | 0 | 0 | 0 |
| IC | 126 | BeEN | 100 | 0 | 100 | 0 | 0 | 100 | 0 | 0 | 0 |
| IC | 94 / 32 | BSR | 100 | 0 | 100 | 60 | 0 | 30 | 20 | 20 | 20 |
| IC | 94 / 32 | BeEN | 100 | 0 | 100 | 0 | 0 | 100 | 0 | 0 | 0 |

**Legend table 3:** Per cohort size and implemented regression methodology the selection probability of each considered predictor is shown in %.
For cohort sizes without a testing/training split these represent the results of one analysis; For cohort sizes with testing/training split these represent the average inclusion frequencies among 10 randomly resampled analyses.
\* Cohort: FC (Full cohort), IC (Intervention cohort)
\*\* Cohort sizes: Cohort sizes are reported as one number (no subsets) or respective the sizes of the training and testing subsets (training/testing)
\*\*\* Method: BSR (Backwards Stepwise Regression), BeEN (Bootstrap enhanced Elastic-Net)
\*\*\*\* ePAL values based on log-transformed ePAL measurements; VR: Variant Repeats; SI: Somatic Instability

**Table 4:** Performance comparison between BSR and BeEN based regression models.

| Cohort* | Cohort sizes** | Method*** | IS a-Rsq. | IS RMSE | OS a-Rsq. | OS RMSE |
| --- | --- | --- | --- | --- | --- | --- |
| FC | 246 | BSR | 0.230 | 15.060 | . | . |
| FC | 246 | BeEN | 0.220 | 15.150 | . | . |
| FC | 184 / 62 | BSR | 0.242 | 14.946 | -0.008 | 15.829 |
| FC | 184 / 62 | BeEN | 0.217 | 15.277 | 0.087 | 15.423 |
| IC | 126 | BSR | 0.160 | 15.700 | . | . |
| IC | 126 | BeEN | 0.140 | 15.880 | . | . |
| IC | 94 / 32 | BSR | 0.155 | 15.644 | -0.249 | 17.048 |
| IC | 94 / 32 | BeEN | 0.135 | 15.863 | -0.061 | 16.211 |

**Legend table 4:** Statistical performance measures (a-Rsq; RMSE) of linear regression models based on variables selected by either BeEN or BSR. For cohort sizes without a training/testing split these represent the results of a single analysis, for cohort sizes with testing/training splits these represent the average performance measures among 10 randomly resampled analyses. In addition to model-derived performance measures the out-of-sample performance measures were calculated if a training/testing split has been used.
\* Cohort: FC (Full cohort), IC (Intervention cohort)
\*\* Cohort sizes: Cohort sizes are reported as one number (no subsets) or respective the sizes of the training and testing subsets (training/testing)
\*\*\* Method: BSR (Backwards Stepwise Regression), BeEN (Bootstrap enhanced Elastic-Net)
\*\*\*\* IS (In-Sample), OS(Out-of-Sample); a-Rsq (adjusted R-squared), RMSE (Root-Mean-Squared-Error)

In order to study the external validity and the impact of overfitting, the full patient cohort was split into training (n=184, 75% cases) and testing (n=62, 25% cases) subsets. Variable selection and regression model fitting were done using the training sets and evaluated using in-sample (IS) performance measures. These models were then used to predict the results of the testing sets. Out-of-sample (OS) performance measures were obtained by evaluating the accuracy of the predicted testing set results. A much lower OS than IS performance is a sign of overfitting, which is a common phenomenon when working with relatively small sample sizes. To account for random sampling effects, the split between training and testing subsets was repeated 10 times. Among the 10 training datasets, BeEN regression robustly selected the variables ePAL, Age and the interaction term Age*ePAL. Only Variant Repeats was less frequently selected (70%). On the contrary, BSR yielded more variable results in this down sampling experiment, with now also selecting the interaction terms ePAL*SI, Age*SI and Age*ePAL*SI in 50% of the subsets, while SI (70%) and the interaction term Age*ePAL (80%) were not always selected (Table 3). Irrespective of the differences in variable selection, the average in-sample (IS) performance measures were similar between the two approaches and similar to the results obtained by utilizing the full dataset (Table 4). Furthermore, the IS performance measures showed small sampling dependent variation (Fig 2). In contrast, the OS-performance estimates showed large sampling dependent variation and much lower average performance measures compared to the IS-measures for both approaches (a-Rsq: -0.01, 0.09; RMSE: 15.83, 15,42; for respectively the BSR and BeEN based regression models; Table 4 and Fig 2). To further study the impact of sample size, all models were run on the intervention cohort only (n=126). As expected, performance further decreased and BSR showed more variability in the predictors selected than BeEN regression (Table 4 and Fig 2).

**Figure 2:**
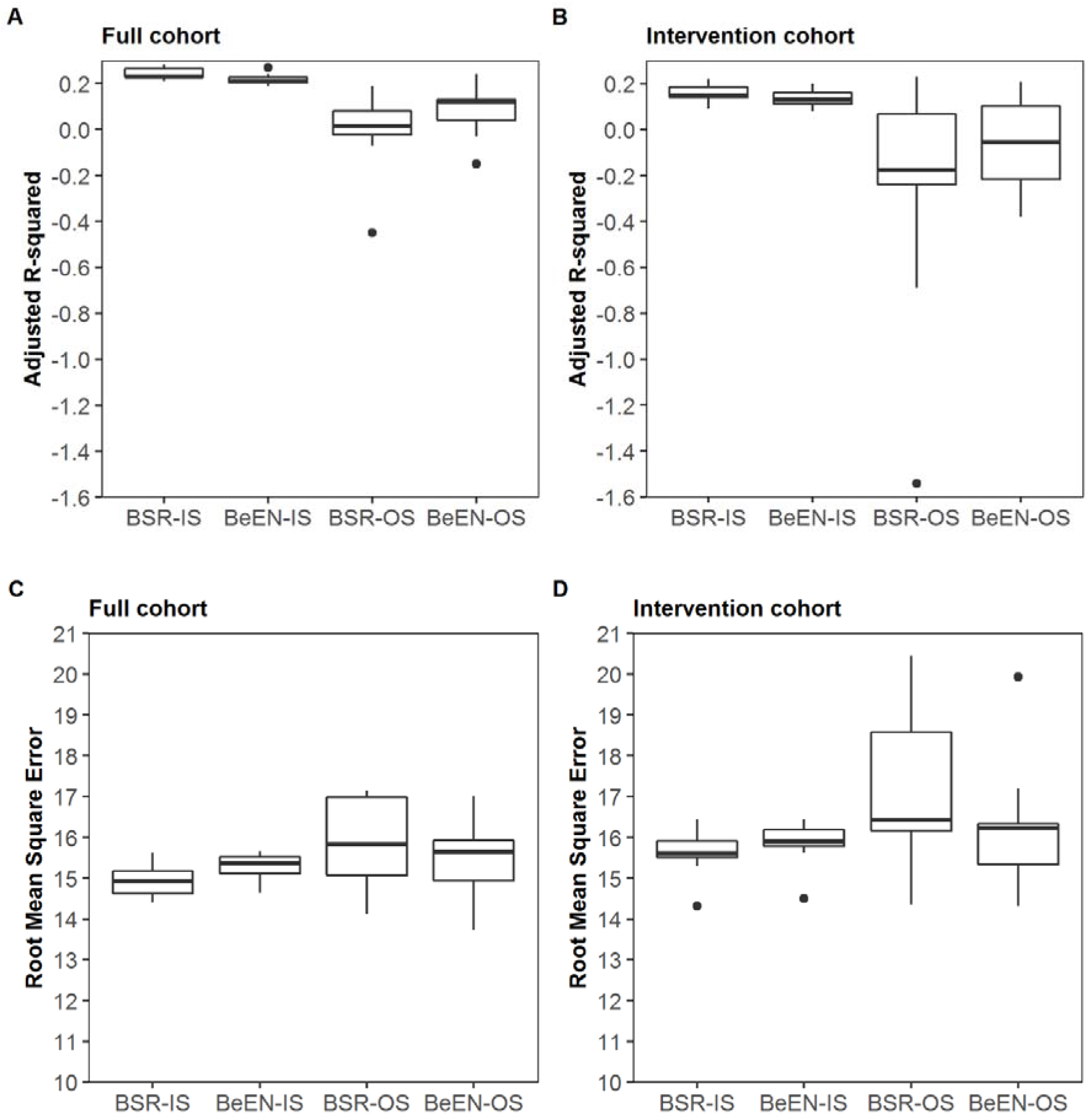
Sampling dependent impact on statistical performance for BSR and BeEN based regression models. Box-plots illustrating the sampling effects on in-sample (IS) and out-of-sample (OS) statistical measures (Adjusted R-squared (panel **A, B**) and Root-Mean-Squared-Error (panel C,D)) when using 10 different 75% training / 25% testing splits of the full cohort (panels **A** and **C**, N=184/62) and the Intervention cohort (panels **B** and **D**, N=94/32).

### Delta-DM1-Activ-c prediction

After BeEN regression framework validation, we subsequently used BeEN regression to select variables associated with response to the behavioural intervention. As penalized regression models necessitate complete datasets, missing values were imputed using Multiple Imputation by Chained Equations (MICE)(23,25). To account for the effects of imputation, 10 different imputed datasets were generated.

Variable inclusion probabilities (VIP) for the potential predictors of delta-DM1-Activ-c are shown in Table 5 for each imputed dataset. For the majority of the predictors stable, imputation independent VIPs were observed. The variables that have been selected in more than 60% of the imputed datasets are most likely the best and most robust prediction variables. These were DM1-Activ-c, SSLI, MIRS, Sex, nIndications, TMT, Stroop, SSLD and BDIFS. The largest variability in VIPs was observed for the variables with the most missing values (MeanENMO, L5ENMO, M5ENMO, ASBQ, SSLD).

**Table 5:**
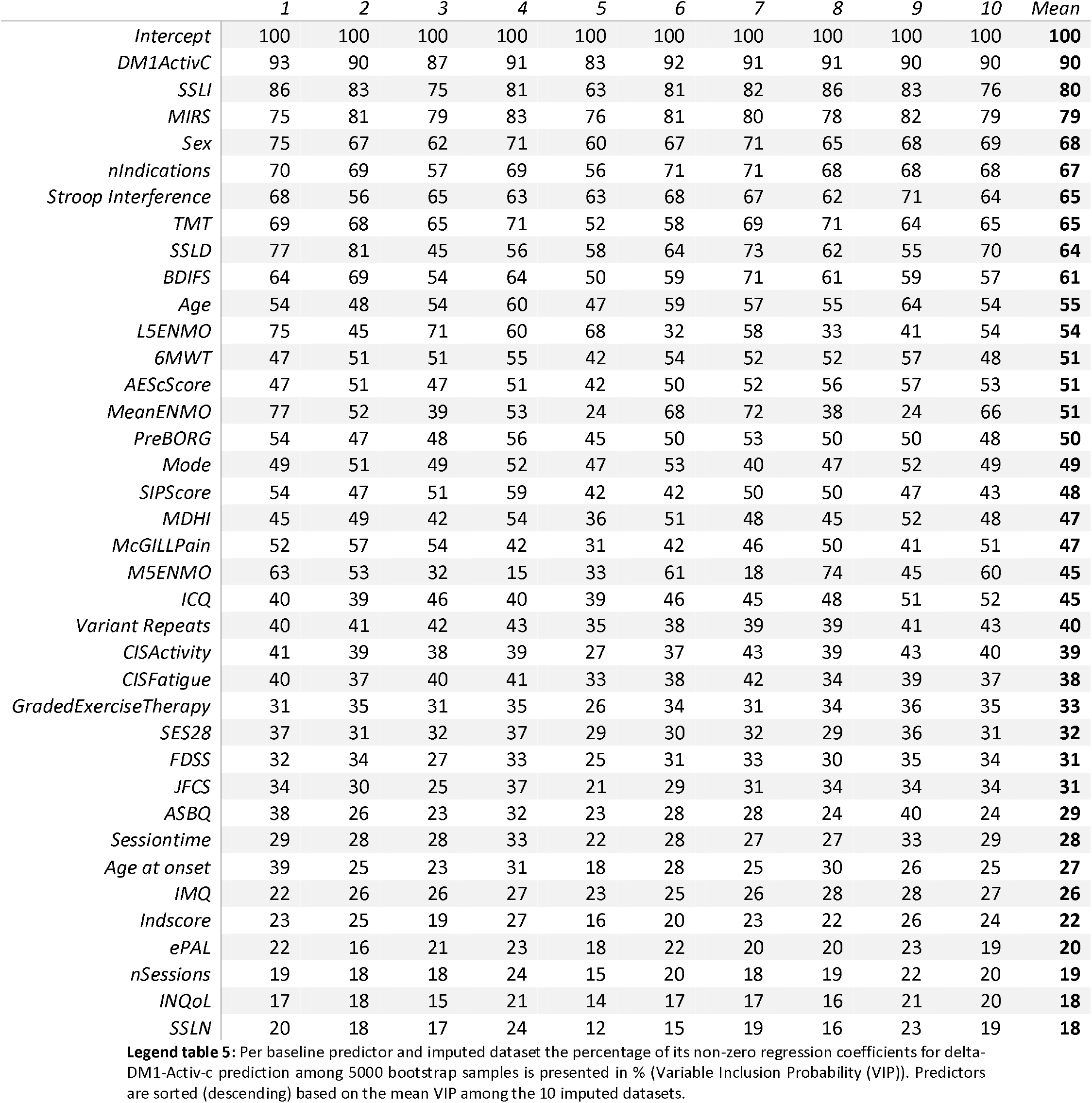
Variable Inclusion Probabilities per imputed dataset.

|  | 1 | 2 | 3 | 4 | 5 | 6 | 7 | 8 | 9 | 10 | Mean |
| --- | --- | --- | --- | --- | --- | --- | --- | --- | --- | --- | --- |
| *Intercept* | 100 | 100 | 100 | 100 | 100 | 100 | 100 | 100 | 100 | 100 | **100** |
| *DM1ActivC* | 93 | 90 | 87 | 91 | 83 | 92 | 91 | 91 | 90 | 90 | **90** |
| *SSLI* | 86 | 83 | 75 | 81 | 63 | 81 | 82 | 86 | 83 | 76 | **80** |
| *MIRS* | 75 | 81 | 79 | 83 | 76 | 81 | 80 | 78 | 82 | 79 | **79** |
| *Sex* | 75 | 67 | 62 | 71 | 60 | 67 | 71 | 65 | 68 | 69 | **68** |
| *nIndications* | 70 | 69 | 57 | 69 | 56 | 71 | 71 | 68 | 68 | 68 | **67** |
| *Stroop Interference* | 68 | 56 | 65 | 63 | 63 | 68 | 67 | 62 | 71 | 64 | **65** |
| *TMT* | 69 | 68 | 65 | 71 | 52 | 58 | 69 | 71 | 64 | 65 | **65** |
| *SSLD* | 77 | 81 | 45 | 56 | 58 | 64 | 73 | 62 | 55 | 70 | **64** |
| *BDIFS* | 64 | 69 | 54 | 64 | 50 | 59 | 71 | 61 | 59 | 57 | **61** |
| *Age* | 54 | 48 | 54 | 60 | 47 | 59 | 57 | 55 | 64 | 54 | **55** |
| *L5ENMO* | 75 | 45 | 71 | 60 | 68 | 32 | 58 | 33 | 41 | 54 | **54** |
| *6MWT* | 47 | 51 | 51 | 55 | 42 | 54 | 52 | 52 | 57 | 48 | **51** |
| *AEScScore* | 47 | 51 | 47 | 51 | 42 | 50 | 52 | 56 | 57 | 53 | **51** |
| *MeanENMO* | 77 | 52 | 39 | 53 | 24 | 68 | 72 | 38 | 24 | 66 | **51** |
| *PreBORG* | 54 | 47 | 48 | 56 | 45 | 50 | 53 | 50 | 50 | 48 | **50** |
| *Mode* | 49 | 51 | 49 | 52 | 47 | 53 | 40 | 47 | 52 | 49 | **49** |
| *SIPScore* | 54 | 47 | 51 | 59 | 42 | 42 | 50 | 50 | 47 | 43 | **48** |
| *MDHI* | 45 | 49 | 42 | 54 | 36 | 51 | 48 | 45 | 52 | 48 | **47** |
| *McGILLPain* | 52 | 57 | 54 | 42 | 31 | 42 | 46 | 50 | 41 | 51 | **47** |
| *M5ENMO* | 63 | 53 | 32 | 15 | 33 | 61 | 18 | 74 | 45 | 60 | **45** |
| *ICQ* | 40 | 39 | 46 | 40 | 39 | 46 | 45 | 48 | 51 | 52 | **45** |
| *Variant Repeats* | 40 | 41 | 42 | 43 | 35 | 38 | 39 | 39 | 41 | 43 | **40** |
| *CISActivity* | 41 | 39 | 38 | 39 | 27 | 37 | 43 | 39 | 43 | 40 | **39** |
| *CISFatigue* | 40 | 37 | 40 | 41 | 33 | 38 | 42 | 34 | 39 | 37 | **38** |
| *GradedExerciseTherapy* | 31 | 35 | 31 | 35 | 26 | 34 | 31 | 34 | 36 | 35 | **33** |
| *SES28* | 37 | 31 | 32 | 37 | 29 | 30 | 32 | 29 | 36 | 31 | **32** |
| *FDSS* | 32 | 34 | 27 | 33 | 25 | 31 | 33 | 30 | 35 | 34 | **31** |
| *JFCS* | 34 | 30 | 25 | 37 | 21 | 29 | 31 | 34 | 34 | 34 | **31** |
| *ASBQ* | 38 | 26 | 23 | 32 | 23 | 28 | 28 | 24 | 40 | 24 | **29** |
| *Sessiontime* | 29 | 28 | 28 | 33 | 22 | 28 | 27 | 27 | 33 | 29 | **28** |
| *Age at onset* | 39 | 25 | 23 | 31 | 18 | 28 | 25 | 30 | 26 | 25 | **27** |
| *IMQ* | 22 | 26 | 26 | 27 | 23 | 25 | 26 | 28 | 28 | 27 | **26** |
| *Indscore* | 23 | 25 | 19 | 27 | 16 | 20 | 23 | 22 | 26 | 24 | **22** |
| *ePAL* | 22 | 16 | 21 | 23 | 18 | 22 | 20 | 20 | 23 | 19 | **20** |
| *nSessions* | 19 | 18 | 18 | 24 | 15 | 20 | 18 | 19 | 22 | 20 | **19** |
| *INQoL* | 17 | 18 | 15 | 21 | 14 | 17 | 17 | 16 | 21 | 20 | **18** |
| *SSLN* | 20 | 18 | 17 | 24 | 12 | 15 | 19 | 16 | 23 | 19 | **18** |

**Legend table 5:** Per baseline predictor and imputed dataset the percentage of its non-zero regression coefficients for delta-DM1-Activ-c prediction among 5000 bootstrap samples is presented in % (Variable Inclusion Probability (VIP)). Predictors are sorted (descending) based on the mean VIP among the 10 imputed datasets.

In order to obtain coefficient estimates, the most frequently selected variables were used to fit a linear regression model on the unimputed intervention cohort dataset. These results hinted towards positive CBT response if patients are female, have high baseline scores of BDIFS, TMT, SSLI and nIndications, and lower baseline scores of DM1-Activ-c, Stroop, MIRS and SSLD (Table 6).

**Table 6:**
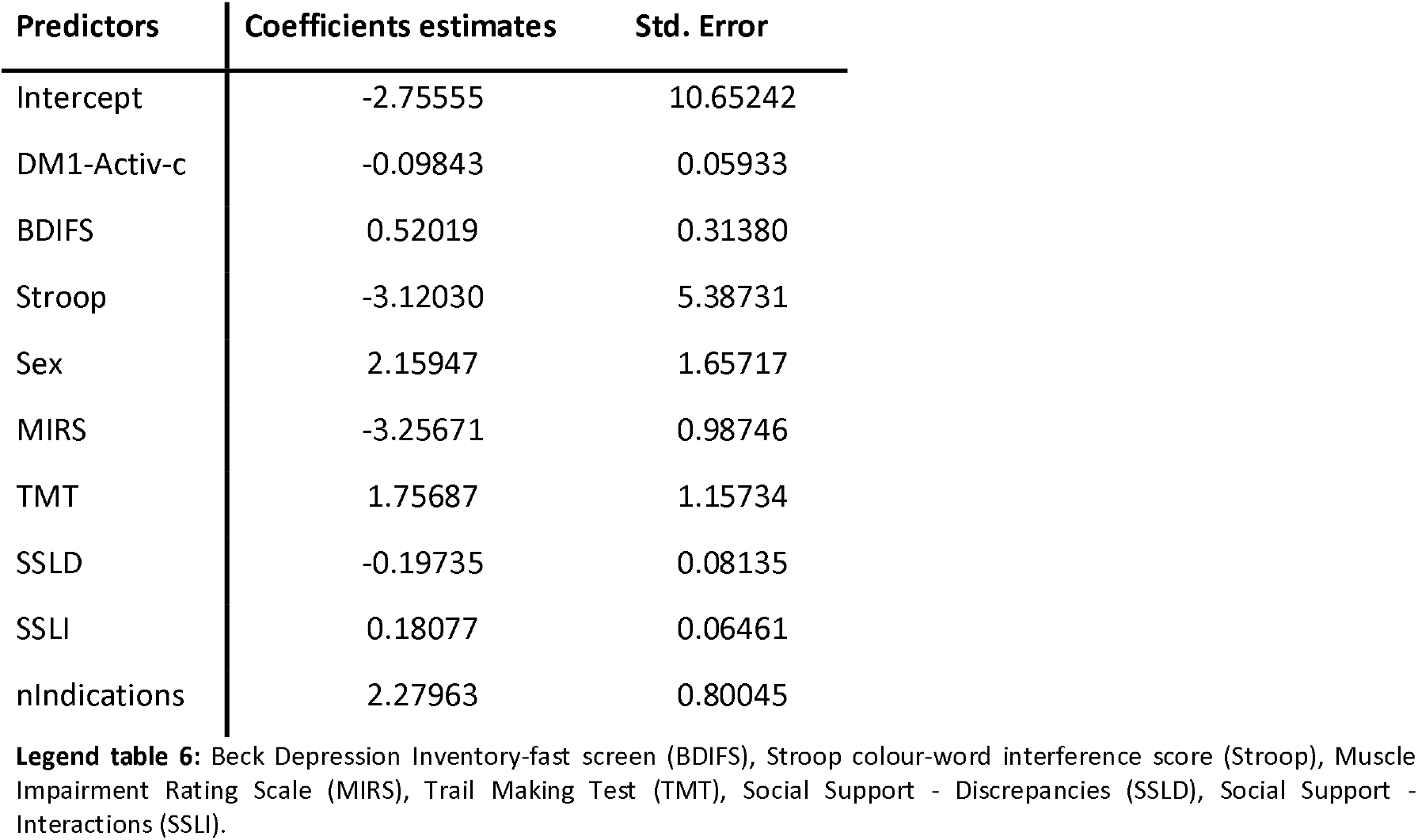
delta-DM1-Activ-c predictor coefficient estimates?

| Predictors | Coefficients estimates | Std. Error |
| --- | --- | --- |
| Intercept | -2.75555 | 10.65242 |
| DM1-Activ-c | -0.09843 | 0.05933 |
| BDIFS | 0.52019 | 0.31380 |
| Stroop | -3.12030 | 5.38731 |
| Sex | 2.15947 | 1.65717 |
| MIRS | -3.25671 | 0.98746 |
| TMT | 1.75687 | 1.15734 |
| SSLD | -0.19735 | 0.08135 |
| SSLI | 0.18077 | 0.06461 |
| nIndications | 2.27963 | 0.80045 |

**Legend table 6:** Beck Depression Inventory-fast screen (BDIFS), Stroop colour-word interference score (Stroop), Muscle Impairment Rating Scale (MIRS), Trail Making Test (TMT), Social Support - Discrepancies (SSLD), Social Support - Interactions (SSLI).

The same regression framework was used to predict delta-DM1-Activ-c in the control (no intervention) group. Here, no predictors with VIPs higher than 60% were identified, suggesting that the predictors in the intervention group are more likely modelling the intervention response and not the natural disease progression.

## Discussion

In this study, we evaluated (1) correlations between cross-sectional data of reported outcome measures, clinical tests and accelerometery derived from the European OPTIMISTIC trial in myotonic dystrophy type 1 (DM1); (2) correlations between longitudinal changes in these outcome measures in the same study(10). Next (3), we validated the Bootstrap enhanced Elastic-Net (BeEN) regression methodology as a valuable tool to select predictors of a clinical intervention with robust outcomes in small cohorts. Last (4), we used the validated BeEN regression methodology to identify promising predictors of a positive effect of cognitive behavioural therapy (CBT) for DM1 patients. In the following paragraphs, we will discuss our results in this order.

In the cross-sectional analysis, OPTIMISTIC’s primary outcome variable, the DM1-Activ-c, demonstrated moderate to strong correlations with measures for exercise capacity (6MWT), objectively assessed activity (MeanENMO, M5ENMO), disease impact (MDHI), quality of life (INQoL) and disease severity (MIRS). In addition, significant yet weaker correlations of DM1-Activ-c scores were seen for several measures of fatigue and fatigue related believes (CIS-fatigue, JFCS, SES28) and with patient reported activity (CIS-activity). However, DM1-Activ-c failed to correlate significantly with measurements of perceived social support (SSLD, SSLI, SSLN) and had only very weak to non-existent correlations with apathy measures (AESc, AESI). This is a noteworthy drawback, as patients with DM1 are severely limited by apathy and decreased social functioning.

Overall, these results support the utility of DM1-Activ-c as a patient reported outcome measure (PROM). If used as a cross-sectional instrument, DM1-Activ-c provides assessments of a variety of important disease aspects. Yet, the questionnaire is not entirely comprehensive, as some of the important disease characteristics are not or only marginally captured. Given the complex multisystem nature of DM1, it may not be surprising that one questionnaire is unable to comprehensively assess all affected disease domains. Nonetheless, these findings can aid in the selection of fewer clinical outcomes in future trials, thereby reducing patient burden while improving study efficiency.

Our longitudinal analysis focused at the changes of outcome parameters over the course of the trial and evaluated whether the different outcome measures changed similarly over time. In general, weaker correlations were found in the longitudinal analyses in comparison to the correlations observed in the cross-sectional analyses, including those for DM1-Activ-c with other variables. Yet, in line with the baseline correlations, delta DM1-Activ-c (dDM1-Activ-c) scores still showed significant, yet weaker correlations with delta scores for measures of objectively assessed activity (dMeanENMO), disease impact (dMDHI), quality of life (dINQoL), patient reported activity (dCIS-activity) and perceived fatigue (dJFCS). Notably, the moderately-strong baseline correlation between DM1-Activ-c and 6MWT scores was not found when analysing delta scores.

The weak longitudinal correlations highlight the heterogeneity in clinical responses towards CBT. To a certain extent this was in line with the expectation, as OPTIMISTIC’s CBT intervention was specifically tailored towards the needs of individual patients. Nonetheless, the absent longitudinal correlations of delta-DM1-Activ-c and delta-6MWT was surprising as both scores improved significantly in the intervention group. These results suggest that improvement on activity and social participation may be independent of gains in exercise capacity. Similar response heterogeneity has been observed in earlier studies, in which improved physical abilities and increased daily activity levels did not mediate fatigue(26). Irrespective of the clinical intervention effects, these results highlight the necessity to choose trial specific outcome measures, as no single outcome measure was able to comprehensively assess the heterogenous intervention effects.

The results of the regression frame work validation suggest that Bootstrap enhanced Elastic-Net (BeEN) regression and Backwards Stepwise Regression (BSR) are mostly selecting consistent predictors. Overall, BeEN regression appears to be a more robust variable selection method, in particular for smaller sample sizes. Independent of cohort sizes and variable selection method applied, the variables ePAL and Age have always been selected, confirming previously established negative associations between variant repeat length and age and disease severity(19,27). Given the comparable IS-performance estimates among the regression models based on either variable selection approach, these variables likely have the strongest predictive effect on baseline DM1-Activ-c among the subset of predictors considered.

However, irrespective of the different down sampling based cohorts used, substantial, methodology independent, variance among the out-of-sample (OS) performance measures has been found. These results hint towards an insufficiently large cohort size to allow for training/testing splits in which both accurately represent the study population. Based on these findings it was concluded that the intervention dataset was sufficiently large to identify variable associated with treatment response when using BeEN regression, but too small to also create a linear model and test its OS statistical performance.

The final aim of this study was to identify variables associated with the response to the CBT intervention. Using BeEN regression, we robustly selected 9 variables that could potentially be associated with the response of the intervention: baseline DM1-Activ-c, BDIFS, Stroop tests, Sex, MIRS, Trail Making Test, SSLD, SSLI and the number of indicated CBT modules. Unsurprisingly, the DM1-Activ-c baseline score was the strongest candidate with a mean Variable Inclusion Probability (VIP) of 90%. This finding may be regarded as a control variable, as potentially positive CBT effects are limited in patients with high baseline scores. The regression estimates of the variables SSLI and SSLD are suggestive of a potential positive effect of experienced social support on the benefits imparted by CBT. Also, the positive regression coefficient of BDIFS implies that patients with more depressive symptoms benefit more from the intervention. This conforms to expectations, as CBT is known to be an effective therapy for depressive disorder(28), aspects of which are present in DM1. Together these findings further support the validity of both the implemented methodology as well as the results. An important potential candidate was the number of CBT modules that were indicated according to baseline screening. Given its positive regression coefficient, this finding implies a positive association between a higher number of indicated modules and intervention effect. This is not surprising, as the DM1-Activ-c scale assesses a broad variety of disease domains, and as such patients are more likely to significantly improve on the DM1-Activ-c scales if several disease domains are addressed. In line with the subgroup analysis of the OPTIMISTIC clinical trial, graded exercise therapy has not been identified as an important predictor of CBT intervention effect(10). The regression coefficients of the final regression model based on the 9 selected predictors as identified by using BeEN regression could provide an indication of directional association of the selected variables with CBT treatment response. However, as the model was built on the same data that has been used to select variables, in-sample statistical estimates cannot give an accurate estimate of the true predictive power of these variables. For this reason, it was decided to not report the associated statistical performance measures of the final model.

## Conclusions

The Rasch-built DM1-Activ-c patient reported outcome scale appears to be a well suited instrument to serve as a marker for a variety of relevant clinical dimensions in DM1. Yet, it is not entirely comprehensive as important disease characteristics such as apathy and certain social measures are not sufficiently captured. Nonetheless, it may make other measurements for exercise capacity, disease impact, quality of life and disease severity redundant if used in a cross-sectional setting. The same does not apply in longitudinal settings, where the correlations between delta scores of DM1-Activ-c with other delta variables were much weaker. As such, researchers are advised to carefully select study-specific outcome measures when designing clinical trials and other longitudinal studies.

Furthermore, we have shown that Bootstrap enhanced Elastic-Net is a robust predictor selection algorithm that can overcome challenges of multidimensionality and multicollinearity. The continuous strive towards personalised medicine is frequently challenged by small patient cohort sizes, especially for rare neuromuscular disorders. Our findings suggest that this machine learning approach is particularly useful for studies with small sample sizes that are typical for clinical trials in rare diseases. Using this approach, a total of 9 predictors were selected that are promising candidates for CBT treatment response prediction. The combined results can guide the design and analysis of prospective trials in DM1 and potentially other neuromuscular disorders.

## Data Availability

Researches whishing to access the OPTIMISTIC trial data should contact BGMvE

## References

1. Fu YH, Pizzuti A, Fenwick RG, King J, Rajnarayan S, Dunne PW, et al. An unstable triplet repeat in a gene related to myotonic muscular dystrophy. Science (80-). 1992;255(5049):1256–8.

2. Brook JD, McCurrach ME, Harley HG, Buckler AJ, Church D, Aburatani H, et al. Molecular basis of myotonic dystrophy: Expansion of a trinucleotide (CTG) repeat at the 31 end of a transcript encoding a protein kinase family member. Cell. 1992 Feb 21;68(4):799–808.

3. Antonini G, Soscia F, Giubilei F, De Carolis A, Gragnani F, Morino S, et al. Health-related quality of life in myotonic dystrophy type 1 and its relationship with cognitive and emotional functioning. J Rehabil Med. 2006 May;38(3):181–5.

4. Kierkegaard M, Harms-Ringdahl K, Holmqvist LW, Tollbck A. Functioning and disability in adults with myotonic dystrophy type 1. Disabil Rehabil. 2011;33(19–20):1826–36.

5. Landfeldt E, Nikolenko N, Jimenez-Moreno C, Cumming S, Monckton DG, Gorman G, et al. Disease burden of myotonic dystrophy type 1. J Neurol. 2019 Apr 4;266(4):998–1006.

6. Minis MAH, Kalkman SJ, Akkermans RP, Engels JA, Huijbregts PA, Bleijenberg G, et al. Employment status of patients with neuromuscular diseases in relation to personal factors, fatigue and health status: A secondary analysis. J Rehabil Med. 2010 Jan;42(1):60–5.

7. Gagnon C, Meola G, Hébert LJ, Puymirat J, Laberge L, Leone M. Report of the first Outcome Measures in Myotonic Dystrophy type 1 (OMMYD-1) international workshop. Clearwater, Florida, November 30, 2011. Neuromuscul Disord. 2013 Dec;23(12):1056–68.

8. Gagnon C, Meola G, Hébert LJ, Laberge L, Leone M, Heatwole C. Report of the second Outcome Measures in Myotonic Dystrophy type 1 (OMMYD-2) international workshop San Sebastian, Spain, October 16, 2013. Neuromuscul Disord. 2015 Jul 1;25(7):603–16.

9. van Engelen B, Abghari S, Aschrafi A, Bouman S, Cornelissen Y, Glennon J, et al. Cognitive behaviour therapy plus aerobic exercise training to increase activity in patients with myotonic dystrophy type 1 (DM1) compared to usual care (OPTIMISTIC): Study protocol for randomised controlled trial. Trials. 2015 May 23;16(1).

10. Okkersen K, Jimenez-Moreno C, Wenninger S, Daidj F, Glennon J, Cumming S, et al. Cognitive behavioural therapy with optional graded exercise therapy in patients with severe fatigue with myotonic dystrophy type 1: a multicentre, single-blind, randomised trial. Lancet Neurol. 2018 Aug 1;17(8):671–80.

11. Hermans MCE, Faber CG, De Baets MH, de Die-Smulders CEM, Merkies ISJ. Rasch-built myotonic dystrophy type 1 activity and participation scale (DM1-Activ). Neuromuscul Disord. 2010 May;20(5):310–8.

12. Hermans MCE, Hoeijmakers JGJ, Faber CG, Merkies ISJ. Reconstructing the Rasch-built myotonic dystrophy type 1 activity and participation scale. PLoS One. 2015 Oct 20;10(10).

13. Sirimongkolkasem T, Drikvandi R. On Regularisation Methods for Analysis of High Dimensional Data. Ann Data Sci. 2019 Dec 13;6(4):737–63.

14. Abram S V., Helwig NE, Moodie CA, DeYoung CG, MacDonald AW, Waller NG. Bootstrap enhanced penalized regression for variable selection with neuroimaging data. Front Neurosci. 2016;10(JUL).

15. Bunea F, She Y, Ombao H, Gongvatana A, Devlin K, Cohen R. Penalized least squares regression methods and applications to neuroimaging. Neuroimage. 2011 Apr 15;55(4):1519–27.

16. Revelle W. Psych: Procedures for Personality and Psychological Research. Illinois; 2018.

17. Wei T, Simko V. R package “corrplot”: Visualization of a Correlation Matrix. 2017.

18. Heymans MW, Van Buuren S, Knol DL, Van Mechelen W, De Vet HCW. Variable selection under multiple imputation using the bootstrap in a prognostic study. BMC Med Res Methodol. 2007 Dec 13;7(1):33.

19. Cumming SA, Jimenez-Moreno C, Okkersen K, Wenninger S, Daidj F, Hogarth F, et al. Genetic determinants of disease severity in the myotonic dystrophy type 1 OPTIMISTIC cohort. Neurology. 2019 Sep 3;93(10):E995–1009.

20. Venables W, Ripley BD. Modern Applied Statistics with S. Fourth Edi. New York: Springer-Verlag New York; 2002.

21. Friedman J, Hastie T, Tibshirani R. Regularization paths for generalized linear models via coordinate descent. J Stat Softw. 2010;33(1):1–22.

22. Canty A, Ripley B. boot: Bootstrap R (S-Plus) Functions. 2017.

23. van Buuren S, Groothuis-Oudshoorn K. mice: Multivariate imputation by chained equations in R. J Stat Softw. 2011 Dec 12;45(3):1–67.

24. Edsel A, Elizabeth H. gvlma: Global validation of Linear Models Assumptions. 2019.

25. Erler NS, Rizopoulos D, Rosmalen J van Jaddoe VWV, Franco OH, Lesaffre EMEH. Dealing with missing covariates in epidemiologic studies: a comparison between multiple imputation and a full Bayesian approach. Stat Med. 2016 Jul 30;35(17):2955–74.

26. Wiborg JF, Knoop H, Stulemeijer M, Prins JB, Bleijenberg G. How does cognitive behaviour therapy reduce fatigue in patients with chronic fatigue syndrome? the role of physical activity. Psychol Med. 2010 Aug;40(8):1281–7.

27. Cumming SA, Hamilton MJ, Robb Y, Gregory H, McWilliam C, Cooper A, et al. De novo repeat interruptions are associated with reduced somatic instability and mild or absent clinical features in myotonic dystrophy type 1. Eur J Hum Genet. 2018 Nov 1;26(11):1635–47.

28. Bagby RM, Quilty LC, Segal Z V, McBride CC, Kennedy SH, Costa PT. Personality and differential treatment response in major depression: a randomized controlled trial comparing cognitive-behavioural therapy and pharmacotherapy. Can J Psychiatry. 2008 Jun 1;53(6):361– 70.

29. Kierkegaard M, Tollbäck A. Reliability and feasibility of the six minute walk test in subjects with myotonic dystrophy. Neuromuscul Disord. 2007 Dec;17(11–12):943–9.

30. Borg G. Borg’s Perceived exertion and pain scales. Champaign IL: Human Kinetics; 1998.

31. Heatwole C, Bode R, Johnson NE, Dekdebrun J, Dilek N, Eichinger K, et al. Myotonic dystrophy health index: Correlations with clinical tests and patient function. Muscle and Nerve. 2016 Feb 1;53(2):183–90.

32. Hermans MCE, Merkies ISJ, Laberge L, Blom EW, Tennant A, Faber CG. Fatigue and daytime sleepiness scale in myotonic dystrophy type 1. Muscle and Nerve. 2013 Jan;47(1):89–95.

33. Worm-Smeitink M, Gielissen M, Bloot L, van Laarhoven HWM, van Engelen BGM, van Riel P, et al. The assessment of fatigue: Psychometric qualities and norms for the Checklist individual strength. J Psychosom Res. 2017 Jul 1;98:40–6.

34. Anderson KN, Catt M J.C. Assessment of sleep and circadian rhythm disorders in the very old: the Newcastle 85+ Cohort Studye. Age Ageing. 2014;43(1):57–63.

35. Vincent KA, Carr AJ, Walburn J, Scott DL, Rose MR. Construction and validation of a quality of life questionnaire for neuromuscular disease (INQoL). Neurology. 2007 Mar;68(13):1051–7.

36. Poole H, Bramwell R, Murphy P. The utility of the Beck Depression Inventory Fast Screen (BDI-FS) in a pain clinic population. Eur J Pain. 2009 Sep;13(8):865–9.

37. Marin RS, Biedrzycki RC, Firinciogullari S. Reliability and validity of the apathy evaluation scale. Psychiatry Res. 1991;38(2):143–62.

38. van Dorst M, Okkersen K, Kessels RPC, Meijer FJA, Monckton DG, van Engelen BGM, et al. Structural white matter networks in myotonic dystrophy type 1. NeuroImage Clin. 2019 Jan 1;21.

39. Mathieu J, Boivin H, Meunier D, Gaudreault M, Bégin P. Assessment of a disease-specific muscular impairment rating scale in myotonic dystrophy. Neurology. 2001 Feb 13;56(3):336–40.

40. Heller LJ, Skinner CS, Tomiyama AJ, Epel ES, Hall PA, Allan J, et al. Trail-Making Test. In: Encyclopedia of Behavioral Medicine. Springer New York; 2013. p. 1986–7.

41. Melzack R. The McGill pain questionnaire: from description to measurement. Anesthesiology. 2005;103(1):199–202.

42. Horwitz EH, Schoevers RA, Ketelaars CEJ, Kan CC, Van Lammeren AMDN, Meesters Y, et al. Clinical assessment of ASD in adults using self- and other-report: Psychometric properties and validity of the Adult Social Behavior Questionnaire (ASBQ). Res Autism Spectr Disord. 2016 Apr 1;24:17–28.

43. Sonderen E van Ormel J. Het meten van aspecten van sociale steun en hun relatie met welbevinden. Een onderzoek naar de bruikbaarheid van de SSL-I ende SSL-D. Gedrag Gezond. 1997;4:190–200.

44. Jacobsen PB, Azzarello LM, Hann DM. Relationship of catastrophizing to fatigue severity in woman with breast cancer. Cancer Res Ther Control. 1999;8:155–64.

45. Evers AW, Kraaimaat FW, van Lankveld W, Jongen PJ, Jacobs JW, Bijlsma JW. Beyond Unfavorable Thinking: The Illness Cognition Questionnaire for Chronic Diseases. J Consult Clin Psychol. 2001;69(6):1026–36.

46. Ray C, Weir W, Stewart D, Miller P, Hyde G. Ways of coping with Chronic Fatigue Syndrome: Development of an illness management questionnaire. Soc Sci Med. 1993;37(3):385–91.

47. Cwm Vos-Vromans D, Smeets RJ, Rijnders LJ, Rm Gorrissen R, Pont M, Köke AJ, et al. Cognitive behavioural therapy versus multidisciplinary rehabilitation treatment for patients with chronic fatigue syndrome: study protocol for a randomised controlled trial (FatiGo). 2012.

48. Robinson BC. Validation of a Caregiver Strain Index. J Gerontol. 1983 May 1;38(3):344–8.

49. Guercio BJ, Donovan NJ, Munro CE, Aghjayan SL, Wigman SE, Locascio JJ, et al. The Apathy Evaluation Scale: A Comparison of Subject, Informant, and Clinician Report in Cognitively Normal Elderly and Mild Cognitive Impairment. J Alzheimer’s Dis. 2015 Jul 24;47(2):421–32.

50. Prcic A, Aganovic D, Hadziosmanovic O. Sickness impact profile (SIP) score, a good alternative instrument for measuring quality of life in patients with ileal urinary diversions. Acta Inform Medica. 2013;21(3):160–5.

